# Identification of a shared, common haplotype cosegregating with an *SGCB* c.544A>C mutation in Indian patients affected with sarcoglycanopathy

**DOI:** 10.1101/2021.12.03.21266857

**Authors:** Shamita Sanga, Sudipta Chakraborty, Mainak Bardhan, Atchayaram Nalini, Moulinath Acharya

**Author notes:** Address correspondence to: Moulinath Acharya, National Institute of Biomedical Genomics, P.O: N.S.S, Kalyani, West Bengal, INDIA 741251. ICMR-National Institute of Cholera & Enteric Diseases, Kolkata.

## Abstract

**Background:** Sarcoglycanopathies (SG) is the most frequent form of autosomal recessive limb-girdle muscular dystrophies (LGMD) leading to progressive muscle wasting and weakness, predominantly characterized by limb-girdle weakness. LGMDR4 is caused by mutations in SGCB encoding for the beta-sarcoglycan proteins. In this study, we describe a shared, common haplotype cosegregating in 14 SG cases from 13 unrelated families with the likely pathogenic homozygous mutation c.544A>C (p.Thr182Pro) in SGCB.

**Methods:** The genotypes of five selected markers (rs10009426, rs6824707, rs2271046, rs35414474 and rs17611952) surrounding the c.544A>C (p.Thr182Pro) were extracted from the variant call format (VCF) generated from whole-exome sequencing (WES) of 14 cases and 14 related family members as controls. The linkage data file was constructed and linkage disequilibrium (LD) plots were generated using HaploView to visualize patterns of LD. Further, haplotype reconstructions based on the 6 markers were conducted using PLINK1.9. using the expectation-maximization (EM) algorithm, an iterative method to find maximum likelihood. Subsequently, the R programming language was used to determine and compare plots of the haplotype frequencies and percentages for both groups to infer the risk haplotypes.

**Results:** Four strong LD blocks were identified in control group: rs10009426 to rs6824707 (0.27□kb), rs6824707 to rs2271046 (41.6 kb), rs10009426 to rs2271046 (41.8 kb) and rs35414474 to rs17611952 (0.17 kb) which were absent in the case group. Similarly, a total of nine haplotypes were estimated in cases and controls of which haplotype H1= G, A, T, G, G, T showed significant statistical difference in the frequency between cases and controls. H1 is also observed to cosegregate with c.544A>C (p.Thr182Pro) in the pedigrees of all the cases.

**Conclusion:** The identification of c.544A>C (p.Thr182Pro) mutation in 14 cases from India indicates a probable event of founder effect. Further, the H1 haplotype, cosegregating with this mutation, convincingly sheds light on the recent developments in population genetics allowing insights into demographic and population history. This haplotype can also be used as a genetic marker to screen individuals with genetic susceptibility as carriers and provide genetically informed risk stratification and management in the prevention of SG.

## Introduction

Limb girdle muscular dystrophies (LGMD) are heterogeneous group of disorders leading to progressive muscle wasting and weakness, predominantly characterized by limb girdle weakness (1). It is caused by mutations in 32 genes causing different type of LGMDs (2). Sarcoglycanopathies (SG) are the most frequent form of autosomal recessive LGMD comprising of four subtypes-LGMDR3, LGMDR4, LGMDR5 and LGMDR6 caused by mutations in *SGCA, SGCB, SGCG* and *SGCD* encoding for the alpha-, beta-, delta- and gamma-sarcoglycan proteins respectively (3). Sarcoglycans are tightly bound to each other and form a transmembrane glycoprotein across the cell membrane of skeletal and cardiac muscle fibres (4). Mutation occurring in one gene may result in partial or total deficiency of all the other sarcoglycan protein in the complex thereby leading to the loss of muscle membrane integrity (5).

In India, Srinivas *et al* (6)(7) presented their initial observations on 35 patients with LGMDs. In the following four decades, several studies have accumulated describing clinical phenotypes of LGMDs and diagnosis achieved by immunohistochemistry. Khadilkar et al. (7) analysed the genetic aspects of 18 SG patients from India. Among them, *SGCG* gene mutation (44.4%) was most common followed by *SGCD* (27.77%), *SGCA* (22.22%), and *SGCB* (5.55%). Among *SGCG* mutation, 525delT was encountered in 50% of the cases although haplotype analysis has not been undertaken.

Even though in recent years, genetic analysis of LGMDs have increasingly been undertaken in various parts of India, there are only few genetically confirmed SG patients with one small series and some case reports available. These efforts are very limited in understanding the prevalence pattern of these diseases in our large country with diverse population. Identifying any founder event within sub-population in these diseases will be very beneficial in genetically informed risk stratification and management of prevention of SG. In this study, we describe a common disease-causing haplotype occurring in 14 SG cases from 13 unrelated families carrying the likely pathogenic homozygous mutation c.544A>C (p.Thr182Pro) in *SGCB*. Interestingly, this mutation is detected in all SG patients coming from the southern states of Tamil Nadu, Karnataka and Andhra Pradesh in India.

## Materials and methods

### Samples selected for haplotyping

Our study sample was composed of 28 individuals from 13 unrelated families originating from the southern states of Tamil Nadu, Karnataka and Andhra Pradesh in India. 14 individuals (including a pair of siblings) were affected and homozygous carrier of c.544A>C mutation in *SGCB* gene, 13 individuals were heterozygous carrier and one individual was normal. The male to female ratio was 1.33 (16 males and 12 females). The patients were genetically confirmed ARLGMD patients at the multidisciplinary neuromuscular disorders clinic from a national referral center for neurological disorders in Bangalore, India. The study was approved by the institutional ethics committee (NIMHANS/IEC/2020-21). Written informed consent was obtained from all participants in the study.

### Haplotype analysis

Five single-nucleotide polymorphisms (SNPs) surrounding the c.544A>C were selected as neutral markers for the haplotype reconstruction: rs10009426, rs6824707, rs2271046, rs35414474 and rs17611952. The genotypes for the six selected markers were extracted from the variant call format (VCF) generated from the BAM files provided by Medgenome, Bangalore, India. These SNPs markers were chosen based on their chromosomal position and on their allelic frequencies. They are located on chromosome 4, in the *DCUN1D4* and *SPATA18* genes, outlining a genomic fragment of approximately 238kb from rs10009426 to rs17611952. The distance from rs10009426 to the c.544A>C is about 184060 bp and from rs17611952 to c.544A>C is 54039bp (table 1) (figure 1B).

**Table 1:** Selection of SNPs as neutral markers surrounding the mutant allele

| Gene | Nucleotide variants | SNP ID | Distance to *SGCB* c.544A>C (bp) | Ancstral Allele | MAF (1KGP3-ALL) | MAF (1KGP3-SAS) | MAF (gnomAD-ALL) | MAF (gnomAD-SAS) |
| --- | --- | --- | --- | --- | --- | --- | --- | --- |
| DCUN1D4 | G/A | rs10009426 | (-) 184060 | G | 0.456 (A) | 0.491 (A) | 0.44 (G) | 0.472 (G) |
| DCUN1D4 | A/G | rs6824707 | (-) 183788 | A | 0.458 (G) | 0.491 (G) | 0.436 (A) | 0.471 (A) |
| DCUN1D4 | T/A | rs2271046 | (-) 142161 | T | 0.455 (A) | 0.491 (A) | 0.357 (T) | 0.45 (T) |
| **SGCB** | **T/G** | **rs751427686** | **0** | **T** | **0 (G)** | **0 (G)** | **1.193e-05 (G)** | **9.8e-05 (G)** |
| SPATA18 | C/G | rs35414474 | (+) 53861 | C | 0.158 (G) | 0.299 (G) | 0.127 (G) | 0.277 (G) |
| SPATA18 | A/T | rs17611952 | (+) 54039 | A | 0.157 (T) | 0.3 (T) | 0.125 (T) | 0.276 (T) |

**Figure 1:**
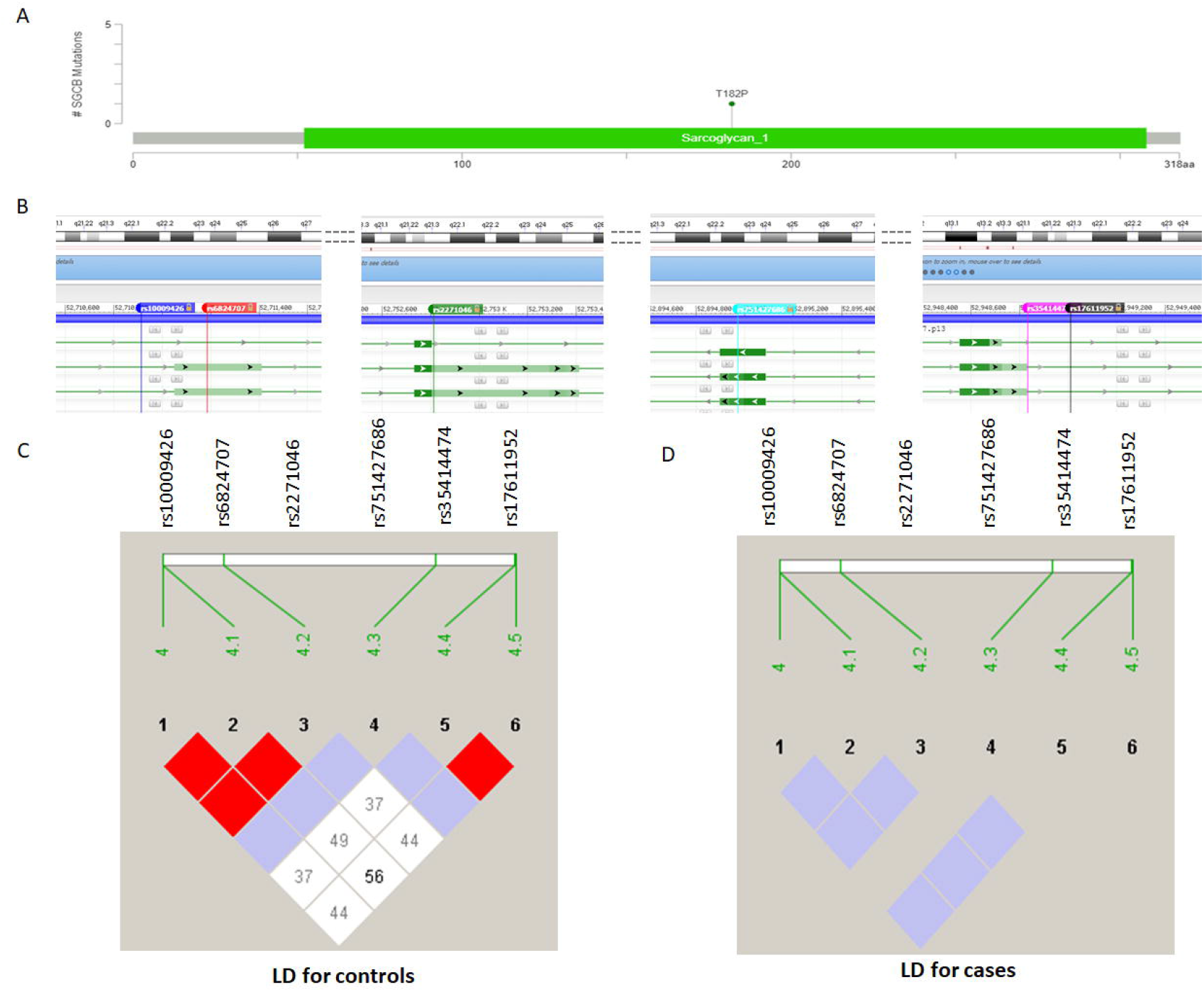
A. Lolliplot to show location of c.544A>C (p.Thr182Pro) mutation in SGCB protein backbone taken from UniProt database. B. Location of the five markers: rs10009426, rs6824707, rs2271046, rs35414474 and rs17611952 surrounding the c.544A>C (p.Thr182Pro) (rs751427686) in the genome data viewer from national centre biotechnology information (NCBI). C. Linkage disequilibrium (LD) plots to visualise patterns of LD for the six selected markers in unaffected controls and D. Affected cases

The linkage data file was constructed including the genotypes of the individuals. Linkage disequilibrium (LD) plots were generated using HaploView (8) to visualise patterns of LD for the two groups (affected cases and unaffected controls) separately to identify the differential pattern of that genic region correlated to disease state. Further, haplotype reconstructions based on the 6 markers selected including the c.544A>C mutation for the two groups were conducted using PLINK1.9 (9). The haplotypes are constructed using multi-marker predictors based on the distribution of probabilistically inferred set of haplotypes for each individual using the expectation–maximization (EM) algorithm, an iterative method to find maximum likelihood. The haplotype frequencies for both cases and controls were estimated. Subsequently, the chi-square test was performed based on the expected number of haplotypes on each individual. A test statistic was computed from genotype that generates sets of haplotype-specific tests with 1 degree of freedom. P-values were generated for each haplotype to observe the significant changes in haplotype distribution for both cases and controls. Subsequently, R programming language was used to determine and compare plots of the haplotype frequencies and percentages for both groups to infer the risk haplotypes.

## Results

The obtained genotypes for all the 28 individuals (cases and controls) for the five selected markers (rs10009426, rs6824707, rs2271046, in the *DCUN1D4* gene and rs35414474 and rs17611952 in the *SPATA18* gene) surrounding c.544A>C (rs751427686) are shown in supplementary table 1. LD plot was constructed in HaploView using the algorithm defined by Gabriel and colleagues (10) to identify strong LD in case and control groups separately. Four strong LD blocks were identified in control group: The first is from rs10009426 to rs6824707 (0.27□kb), second is from rs6824707 to rs2271046 (41.6 kb), third is from rs10009426 to rs2271046 (41.8 kb) and the fourth is from rs35414474 to rs17611952 (0.17 kb) (figure 1C). In the case group, no such LD block was observed (figure 1D).

Haplotype reconstruction based on the 6 markers selected including the c.544A>C mutation for the two groups were conducted using PLINK1.9. A total of nine haplotypes were estimated in cases and controls (figure 2A). On performing chi-square test, significant statistical difference on the frequency of haplotype between cases and controls were observed only for haplotype H1= G, A, T, G, G, T with a P-value of 0.000539. No significant differences were identified for other 8 haplotypes (figure 2B). 13 cases (including a pair of siblings) homozygous for the c.544A>C mutation in 12 unrelated families shared a common homozygous haplotype H1 for all SNPs (rs10009426, rs6824707, rs2271046, rs751427686, rs35414474 and rs17611952) in chromosome 4 (figure 3). However, in one patient (P47, pedigree#11) (figure 3), H1 haplotype is present in one chromosome while a different haplotype (H9= A, G, A, G, G, T) occurs in the other chromosome. The frequency of these two haplotypes H1 and H9 estimated in affected cases (N=14) are 93.75% and 6.25%, respectively (figure 2C).

**Figure 2:**
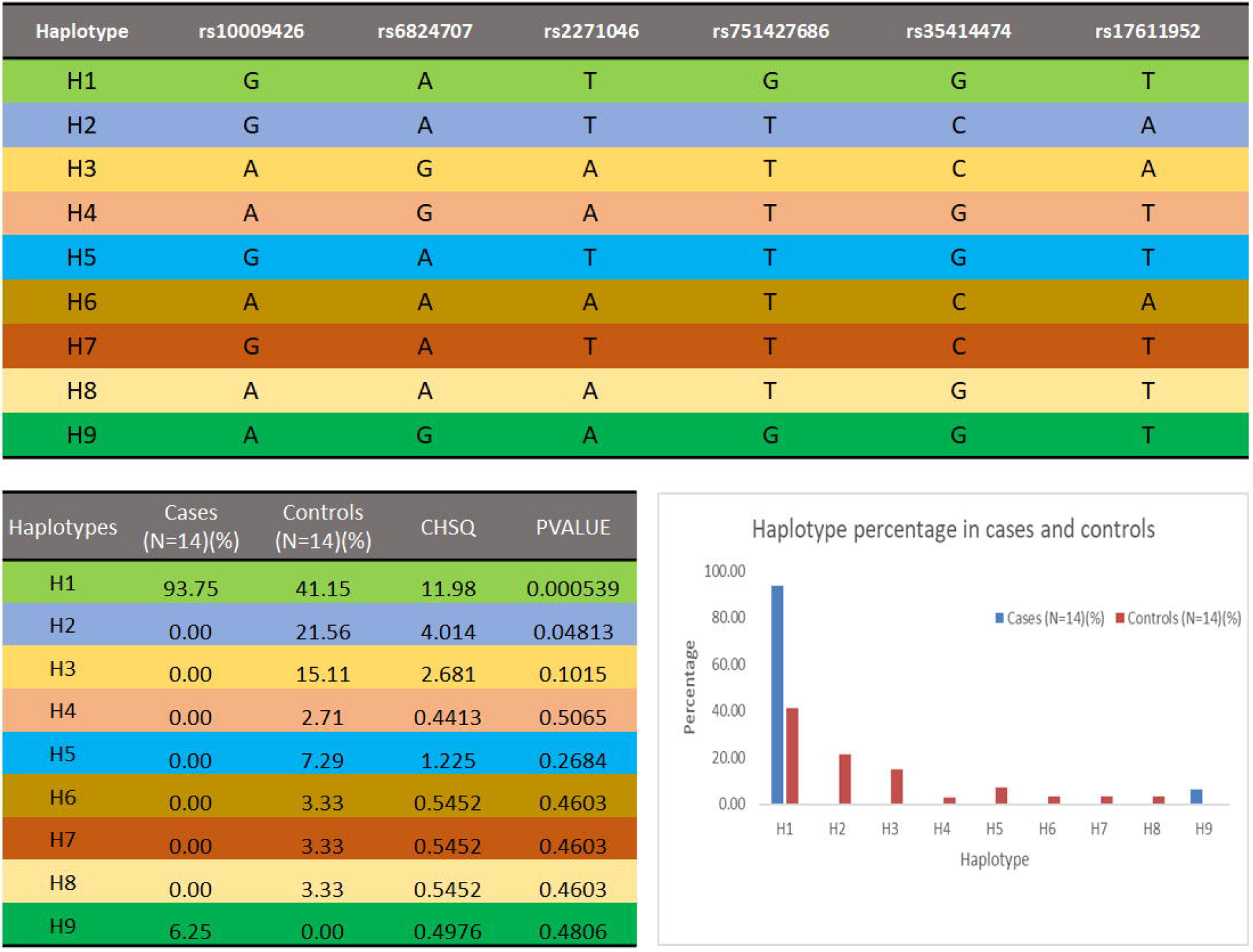
A. The nine haplotypes identified in cases and controls. Each haplotype is colour coded. B. The estimated frequency of the haplotypes in cases and controls. The chi-square (CHSQ) test was performed based on the expected number of haplotypes on each individual. P-values were generated for each haplotype to observe the significant changes in haplotype distribution for both cases and controls. C. Graphical distribution of haplotype percentages in cases and controls.

**Figure 3:**
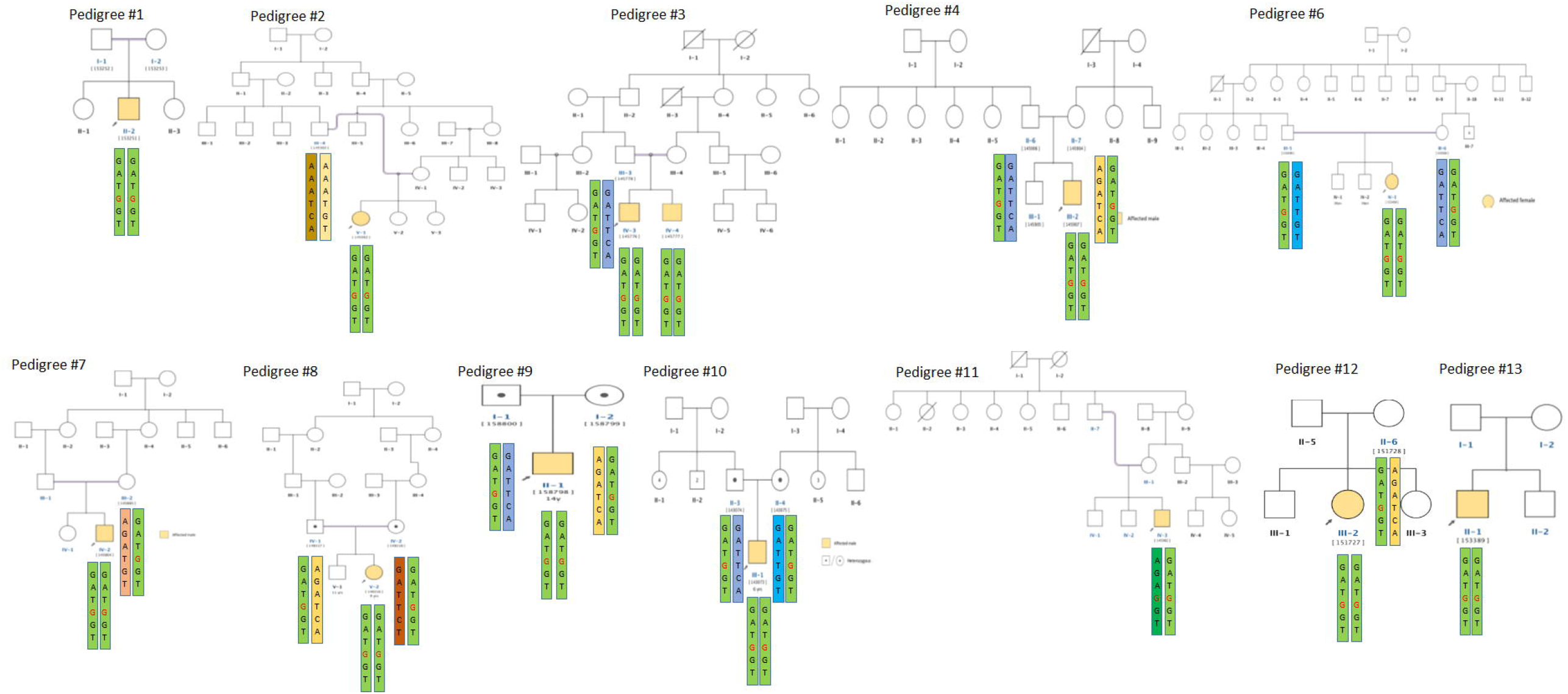
The pedigrees of the 13 cases (including a pair of siblings) showing the distribution of the nine haplotypes identified.

In the related unaffected heterozygous carrier of c.544A>C controls, heterozygous H1 haplotype is present in all controls except one. P37-P1 (parent? of P37 in pedigree#2) (figure 3) is the only normal individual not carrying the c.544A>C mutation and hence the H1 haplotype is absent. The other haplotypes identified in the controls are haplotype H2 (G, A, T, T, C, A) (pedigree#3, 4, 6, 9, 10), H3 (A, G, A, T, C, A) (pedigree#4, 8, 9, 12), H4 (A,G,A,T,G,T) (pedigree#7) H5 (G, A, T, T, G, T) (pedigree#6, 10), H6 (A, A, A, T, C, A) (pedigree#2), H7 (G, A, T, T, C, T) (pedigree#8), H8 (A, A, A, T, G, T) (pedigree#2) and H9 (A, G, A, G, G, T) (pedigree#11) (figure 3) The frequency of these haplotypes estimated in controls (N=14) are H1= 41.15%, H2= 21.56%, H3= 15.11%, H4= 2.71%, H5= 7.29%, H6= 3.33%, H7= 3.33% and H8=3.33% (figure 2C).

## Discussion

We report here a group of Indian patients carrying the c.544A>C mutation in *SGCB* in a homozygous state. This mutation has been previously reported by Ganapathy et al. (11) in two patients one in homozygous state and one in compound heterozygous state with another mutation (c.286G>C) in the *SGCB* gene. This extremely rare variant (c.544A>C, p.Thr182Pro) has been reported in dbSNP (rs751427686), and totally absent in the 1000 genome phase 3 data. However, in the GnomAD database, this rare variant is present with extremely low frequency (1.193e-05 in all and 9.8e-05 in SAS; table 1). The 182^nd^ Threonine residue is located in the extracellular domain of the beta component of Sarcoglycan protein. Changing this to Proline due to the c.544A>C mutation is therefore creating a deleterious and potentially damaging change in the protein, thus making it a likely pathogenic genetic alteration (figure 1A). Furthermore, to the best of our knowledge, this mutation, in the context of sarcoglycanopathies, is not reported in any other population, thereby ruling out the hypothesis that it is caused by a mutation ‘hotspot’. Presence of this mutation in homozygous condition in 13 patients from 12 unrelated families therefore, naturally qualify this as a founder mutation and we wanted to probe further whether the patients harbouring this mutation also share a common haplotype at the *SGCB* locus.

We selected a set of five SNPs as neutral markers (rs10009426, rs6824707, rs2271046, rs35414474 and rs17611952) surrounding the c.544A>C mutation in the *SGCB* gene spanning about 238 kb to visualise patterns of LD for the two groups (affected cases and unaffected controls) separately to identify any possible differential pattern of that genic region correlated to disease state (figure 1B). The markers selected have a high frequency in the 1000 Genome database as well as in the GnomAD (table 1). Interestingly, three SNPs (rs10009426, rs6824707 and rs2271046) located 5’ and two SNPS (rs35414474 and rs17611952) located 3’ to the c.544A>C mutation, were in strong LD with each other in related unaffected controls but completely absent in affected cases (figure 1 C, D). This indicates a possibility of multiple recombination events occurring surrounding the c.544A>C mutation.

This observation encouraged us further to reconstruct and identify a common ancestral haplotype with these five common variants that might be segregating within all the 14 patients from 13 families. We have identified a total of nine haplotypes in both case and control groups arising from these five SNPs and the c.544A>C mutation in *SGCB* gene (figure 2A). Interestingly, haplotype H1 (G, A, T, G, G, T) has been found in all the affected individuals in homozygous condition and segregating within the pedigrees except one. For the patient P47 in pedigree#11, H1 is present with another haplotype H9 (A, G, A, G, G, T), which is totally absent in the unaffected family members (figure 3). The H1 haplotype is also present in all the unaffected carriers in heterozygous condition in combination with other haplotypes (H2, H3, H4, H5, H6, H7 and H8) except H1 and H9. The parent (P37-P1) of the patient P37 in pedigree#2 however, does not share the H1 haplotype in any of his chromosome, instead carrying two completely different haplotypes H6 and H8, making this as a case of doubtful paternity (figure 3). Nonetheless, the homozygous occurrence of the H1 haplotype with clear indication of its segregation pattern in all the families having patients suffering from sarcoglycanopathy designate this as the primary haplotype co-segregating with the c.544A>C mutation in the *SGCB* gene.

In conclusion, to the best of our knowledge, till date no mutation has been identified in Indian patients suffering from sarcoglycanopathy with a possible founder effect. Here we have not only identified a mutation in the *SGCB* gene with a probable founder effect, present in homozygous condition in 13 unrelated patients, but constructed haplotypes with five common variants surrounding the mutation and most importantly, designated a shared, common haplotype H1 segregating completely with the mutation. This could be used as genetic marker to screen individuals with genetic susceptibility as carrier. In addition, a larger sample size for the c.544A>C mutation carriers would help in calculating the age of the most recent common ancestor of all carriers of the mutation. This would shed more light on the recent developments in population genetics allowing insights into demographic and population history.

## Supporting information

Supplemental genotype table

## Data Availability

All data produced in the present study are available upon reasonable request to the authors

## AUTHOR CONTRIBUTIONS

SS, AN and MA conceptualized the work. SS and SC were responsible for generation of LD plots and haplotype construction. Haplotype segregation analyses was done by SS and MA. MB and AN provided with the relevant clinical data. Results were validated by SS, SC and MA. SS, SC and MA designed and wrote the manuscript.

## ACKNOWLEDGEMENTS

The authors thank Medgenome, Bangalore for providing BAM and VCF files for sarcoglycanopathy patients and their unaffected family members. SS is a recipient of a doctoral fellowship from CSIR, India. SC is a recipient of doctoral fellowship from ICMR, India. The authors thank Partha Pratim Majumder for scientific discussion regarding haplotype analyses. The authors also thank intramural support from NIBMG to conduct this work.

