## Supplemental genotype table for "Identification of a shared, common haplotype cosegregating with an *SGCB* c.544A>C mutation in Indian patients affected with sarcoglycanopathy"

Supplementary table: The genotypes of 5 markers selected surrounding the c.544A>C (p.Thr182Pro) (rs751427686) mutation for the cases and unaffected related controls.

| **Pedigree #** | **ID** | **rs10009426** | **rs6824707** | **rs2271046** | **rs751427686** | **rs35414474** | **rs17611952** |
| --- | --- | --- | --- | --- | --- | --- | --- |
| Ped1 | P36 | G/G | A/A | T/T | G/G | G/G | T/T |
| Ped2 | P37 | G/G | A/A | T/T | G/G | G/G | T/T |
| Ped2 | P37-P1 | A/A | A/A | A/A | T/T | C/G | A/T |
| Ped3 | P38 | G/G | A/A | T/T | G/G | G/G | T/T |
| Ped3 | P38-S | G/G | A/A | T/T | G/G | G/G | T/T |
| Ped3 | P38-P1 | G/G | A/A | T/T | G/T | C/G | A/T |
| Ped4 | P39-P1 | G/G | A/A | T/T | G/T | C/G | A/T |
| Ped4 | P39-P2 | A/G | G/A | A/T | G/T | C/G | A/T |
| Ped4 | P39 | G/G | A/A | T/T | G/G | G/G | T/T |
| Ped5 | P41 | G/G | A/A | T/T | G/G | G/G | T/T |
| Ped6 | P40 | G/G | A/A | T/T | G/G | G/G | T/T |
| Ped6 | P40-P1 | G/G | A/A | T/T | G/T | G/G | T/T |
| Ped6 | P40-P2 | G/G | A/A | T/T | G/T | C/G | A/T |
| Ped7 | P43 | G/G | A/A | T/T | G/G | G/G | T/T |
| Ped7 | P43-P2 | A/G | G/A | A/T | G/T | G/G | T/T |
| Ped8 | P44 | G/G | A/A | T/T | G/G | G/G | T/T |
| Ped8 | P44-P1 | A/G | G/A | A/T | G/T | C/G | A/T |
| Ped8 | P44-P2 | G/G | A/A | T/T | G/T | C/G | T/T |
| Ped9 | P45 | G/G | A/A | T/T | G/G | G/G | T/T |
| Ped9 | P45-P2 | G/G | A/A | T/T | G/T | C/G | A/T |
| Ped9 | P45-P1 | A/G | G/A | A/T | G/T | C/G | A/T |
| Ped10 | P46 | G/G | A/A | T/T | G/G | G/G | T/T |
| Ped10 | P46-P1 | G/G | A/A | T/T | G/T | C/G | A/T |
| Ped10 | P46-P2 | G/G | A/A | T/T | G/T | G/G | T/T |
| Ped11 | P47 | A/G | G/A | A/T | G/G | G/G | T/T |
| Ped12 | P48 | G/G | A/A | T/T | G/G | C/G | T/T |
| Ped12 | P48-P2 | A/G | G/A | A/T | G/T | C/G | A/T |
| Ped13 | P51 | G/G | A/A | T/T | G/G | G/G | T/T |
